# An Electronic Health Record Compatible Model to Predict Personalized Treatment Effects from the Diabetes Prevention Program: A Cross-Evidence Synthesis Approach Using Clinical Trial and Real World Data

**DOI:** 10.1101/2021.01.06.21249334

**Authors:** David M Kent, Jason Nelson, Anastassios Pittas, Francis Colangelo, Carolyn Koenig, David van Klaveren, Elizabeth Ciemins, John Cuddeback

## Abstract

**Background:** An intensive lifestyle modification program or metformin pharmacotherapy reduced the risk of developing diabetes in patients at high risk, but are not widely used in the 88 million American adults with prediabetes.

**Objective:** Develop an electronic health record (EHR)-based risk tool that provides point-of-care estimates of diabetes risk to support targeting interventions to patients most likely to benefit.

**Design:** Cross-design synthesis: risk prediction model developed and validated in large observational database, treatment effect estimates from risk-based reanalysis of clinical trial data.

**Setting:** Outpatient clinics in US.

**Patients:** Risk model development cohort: 1.1 million patients with prediabetes from the OptumLabs Data Warehouse (OLDW); validation cohort: distinct sample of 1.1 million patients in OLDW. Randomized clinical trial cohort: 3081 people from the Diabetes Prevention Program (DPP) study.

**Interventions:** Randomization in the DPP: 1) an intensive program of lifestyle modification; 2) standard lifestyle recommendations plus 850 mg metformin twice daily; or 3) standard lifestyle recommendations plus placebo twice daily.

**Results:** Eleven variables reliably obtainable from the EHR were used to predict diabetes risk. This model validated well in the OLDW (*c*-statistic = 0.76; observed 3-year diabetes rate was 1.8% in lowest-risk quarter and 19.6% in highest-risk quarter). In the DPP, the hazard ratio for lifestyle modification was constant across all levels of risk (HR = 0.43, 95% CI 0.35 – 0.53); while the HR for metformin was highly risk-dependent (HR HR = 1.1 [95% CI: 0.61 - 2.0] in the lowest-risk quarter vs. HR=0.45 [95% CI: 0.35 - 0.59] in the highest risk quarter). Fifty-three percent of the benefits of population-wide dissemination of the DPP lifestyle modification, and 76% of the benefits of population-wide metformin therapy can be obtained targeting the highest risk quarter of patients.

**Limitations:** Differences in variable definitions and in missingness across observational and trial settings may introduce estimation error in risk-based treatment effects.

**Conclusion:** An EHR-compatible risk model might support targeted diabetes prevention to more efficiently realize the benefits of the DPP interventions.

## Introduction

The Diabetes Prevention Program (DPP) Study showed that either an intensive program of lifestyle modification or pharmacotherapy with metformin substantially reduced the risk of developing type 2 diabetes in patients at high risk, compared to “usual care.”^1^ The findings have broad implications, as “prediabetes” affects approximately 88 million US adults in the US.^2^

Strenuous calls to address the epidemic of diabetes with prevention^3,4^ have been counter-balanced by concerns about the over-medicalization of prediabetes.^5^ Almost two decades after the publication of the DPP Study, it remains unclear how best to implement these interventions in such an overwhelmingly large, and mostly undiagnosed, population. A 2015 study examining a national sample of over 17,000 working-age adults with prediabetes found that only 3.7% were receiving metformin.^6^ Similarly, widespread use of the intensive lifestyle intervention remains largely unrealized despite evidence that rigorous diet and physical activity promotion reduces diabetes risk in the community setting.^7^

Yet, prediabetes is itself a heterogeneous condition. We previously showed that even among patients enrolled in the DPP Study itself, the risk of developing diabetes within 3 years varies widely and is highly skewed.^8^ Some trial participants were estimated to have a 1–2% risk, others 90%. Unsurprisingly, the degree of benefit from metformin or from the lifestyle intervention was also distributed unevenly.

This prior proof-of-concept work had several limitations. Notably, the risk distribution within the DPP trial participants may differ from that of patients seen in routine practice, particularly since the American Diabetes Association (ADA) has subsequently broadened its definition of prediabetes to include a still more heterogeneous population.^9^ Further, the application of prediction methods to data routinely collected in the electronic health record (EHR) provides a promising means to overcome some of the major barriers to the use of risk models.^10,11^ For example, in addition to requiring manual ascertainment of variables, the previously reported DPP-based model required waist circumference and waist-to-hip ratio measurements that are not difficult to ascertain in routine practice. Herein, we describe development of a clinical prediction model using a hybrid approach that makes use of routinely collected EHR data to predict the risk of diabetes onset and clinical trial data to estimate unbiased risk-based effects of preventive interventions.

## Methods

### Overview

We sought to develop and validate a diabetes risk prediction model using data elements readily available in the EHR for dissemination across healthcare systems as an EHR-embedded tool, to facilitate ease of use. The tool provides clinicians and their patients with an individualized risk of developing diabetes and the estimated benefit of applying a DPP treatment strategy—either an intensive lifestyle program or pharmacotherapy with metformin (the combination of both was not tested in the DPP Study).

### Data sources and Participants

The model was developed and validated using EHR data from the OptumLabs Data Warehouse (OLDW). The OptumLabs EHR database is a geographically diverse sample of the US population with longitudinal clinical data on over 33 million lives with at least one clinic visit during the study period. Using a retrospective observational cohort design, we geographically stratified the database by US Census Region into a development cohort of 1.1 million patients (North East, South, and West) and a separate validation cohort of 1.1 million patients (Midwest).

Eligibility criteria included age between 25 and 75 upon an “index” office or clinic encounter (“index visit” defined by CPT/HCPCS codes, see Appendix Table 1) between January 1, 2012, and December 31, 2016, at which time they met lab-based criteria for the diagnosis of prediabetes. Prediabetes was defined by current American Diabetes Association (ADA) criteria, i.e., having no diagnosis of type 1 or type 2 diabetes on the problem list and one of the following within 12 months prior to the visit: hemoglobin A1c between 5.7 and 6.4% inclusive and/or fasting glucose (FG) between 100 and 125 mg/dL inclusive. Since labeling of fasting status may be incomplete, a glucose drawn at the same time as a lipid panel or triglycerides was considered as fasting. We did not use the 2-hour post glucose load criterion as it is rarely used in clinical practice for prediabetes. Patients were excluded if they had random (non-fasting) glucose greater than or equal to 200 mg/dL on two occasions within a 3-month period prior to the index visit. Women with documented pregnancy within 24 months of the index visit were also excluded. To ascertain development of diabetes, patients also had to have some clinical activity 3 years after the index visit. Eligibility criteria are detailed in Appendix Table 1.

**Table 1.** Cohort Characteristics.

|  |  | OLDW cohort |  |  | DPP |
| --- | --- | --- | --- | --- | --- |
|  |  | Overall | Development | Validation |  |
|  | Missing | n= 2,152,816 | n= 1,076,983 | n= 1,075,833 | n= 3081 |
| Age, years, mean (SD) | 0.0% | 54.9 (11.7) | 55.1 (11.9) | 55 (11.5) | 50.6 (9.0) |
| Female, % | 0.1% | 50.3 | 51.3 | 49.1 | 66.6 |
| Race, % |  |  |  |  |  |
| White | 8.2% | 86.5 | 84.3 | 88.9 | 57.4 |
| Black | 8.2% | 10.2 | 10.8 | 9.1 | 20.9 |
| Other nonwhite race (Optum = Asian) | 8.2% | 3.4 | 4.9 | 1.9 | 5.2 |
| Smoking status, % | 15.6% |  |  |  |  |
| Current smoker |  | 23.3 | 20 | 26.4 | 9.0 |
| Never smoked |  | 48 | 53.2 | 42.9 | 35.2 |
| Former smoker |  | 28.8 | 26.8 | 30.7 | 55.8 |
| Height, cm, mean (SD) | 15.9% | 170.1 (10.1) | 169.5 (10.1) | 170.7 (10) | 166.8 (9.2) |
| Body mass index, mean (SD) | 12.2% | 31.1 (7) | 30.8 (6.7) | 31.8 (6.9) | 33.5 (5.8) |
| Diagnosis of hypertension, n (%) | NA | 44.5 | 44.4 | 45 | 27.1 |
| Systolic blood pressure, mmHg, mean (SD) | 9.0% | 127.4 (14.9) | 127.6 (15.2) | 127.3 (14.7) | 124.2 (14.7) |
| HDL cholesterol, mg/dl, mean (SD) | 12.3% | 50.9 (14.7) | 51.3 (14.9) | 50.6 (14.5) | 45.6 (11.8) |
| Triglycerides, mg/dl, mean (SD) | 12.6% | 138.3 (72.8) | 136.9 (72.8) | 139.7 (72.7) | 162.9 (93.5) |
| Hemoglobin A1c, %, mean (SD) | 54.7% | 5.8 (0.3) | 5.8 (0.3) | 5.8 (0.3) | 5.9 (0.5) |
| Fasting plasma glucose, mg/dl, mean (SD) | 3.8% | 103.7 (10.8) | 103 (11.1) | 104.5 (10.4) | 107.2 (7.7) |
| FPG (fasting), mg/dL | 86.3% | 103.3 (9.2) | 101.3 (10.5) | 105.3 (7.3) |  |
| FPG (random), mg/dL | 13.0% | 103.7 (11.4) | 103.1 (11.4) | 104.4 (11.2) |  |

The DPP dataset was used to estimate treatment effect for metformin or the intensive lifestyle modification program. The design, rationale, outcomes, and loss to follow-up of the DPP have been described in detail elsewhere^1,12^. Briefly, inclusion criteria included a body mass index (BMI) of 24 or higher (22 or higher in Asians) and a fasting plasma glucose concentration of 95 to 125 mg/dL inclusive (impaired fasting glucose) and a concentration of 140 to 199 mg/dL inclusive two hours after a 75 g oral glucose load (impaired glucose tolerance). We note these criteria differ from the ADA’s current diagnostic criteria for prediabetes we used for the OLDW model; the ADA definition imposes no BMI requirement.^13^ The DPP participants were randomized to: 1) standard lifestyle recommendations plus 850 mg of metformin twice daily; 2) an intensive program of lifestyle modification that included 16 lessons with a case manager and set goals of at least a 7 percent weight loss and at least 150 minutes of physical activity per week; or 3) standard lifestyle recommendations plus placebo twice daily. After a median follow-up period of 2.8 (range 1.8–4.6) years, progression to diabetes was reduced by 58% (95% confidence interval, 47% to 66%) in the lifestyle modification arm and 31% (17% to 43%) in the metformin arm, both compared with the placebo arm^1^. The NIDDK Data Repository, from which we obtained data, includes 3081 of the 3234 DPP participants (95% of full population), as some local institutional review boards declined to participate in data distribution.

### Outcome

For the OLDW cohort, the time to event outcome was defined as the time to the first patient encounter after the index visit with documented evidence of type 2 diabetes by any of the following criteria:^14^ diagnosis codes ICD-9 250.x0 or 250.x2 or ICD-10 E11.xx; pharmacotherapy or procedure for type 2 diabetes (as detailed in Appendix Table 1); A1c greater than 6.4%; fasting glucose (or presumed fasting, as noted above) greater than 125 mg/dL; 2-hour OGTT post-load glucose greater than 199 mg/dL. Lab-based criteria required confirmation by an additional lab in the diabetes range or by another method (i.e., diagnosis or medication). Follow-up time for patients who did not meet the outcome definition was censored at the first occurrence of the last observed encounter or end of study period.

### Candidate predictors

A priori risk model predictors were identified by a systematic review conducted by Collins et al.^15^ We selected the following 11 independent variables that were included in at least 3 prior diabetes risk models and were judged to be easily and reliably obtainable in EHR data: age, gender, race, smoking status, BMI, presence or absence of a diagnosis of hypertension, systolic blood pressure, HDL cholesterol, triglycerides, fasting glucose, hemoglobin A1c (HgbA1c). Four variables included in 3 prior models were not considered based on the difficulty of ascertaining them in EHR data: physical activity, waist circumference, waist-to-hip ratio, and family history of diabetes.

### Missing data

Missing data is a common limitation when working with EHR data.^16^ While multiple imputation may improve estimates of parameter effects under a missing-at-random assumption, it does not provide a practical means to cope with missingness in actual patients for whom a prediction needs to be made. Thus, we used missing indicator variables to capture the predictive effects of missingness under the assumption that future and prior missingness are similarly informative. For each predictor, an additional dichotomous variable indicated the presence of missing values. For continuous variables (e.g., BMI, HgbA1c), the missing value of the original variable was replaced by a fixed constant (the median) prior to model estimation, and the missing indicator variable appropriately adjusted for the “missing variable effect.” For categorical variables (e.g., race, smoking status), an additional level was added to define the missing category.

### Model development

We used multivariable Cox proportional hazards regression to estimate the predicted probability of developing type 2 diabetes. We included two a priori interactions, race*BMI and race*HgbA1c, based upon clinical judgment and the literature.^17,18^ Model performance was assessed for discrimination and calibration. A bootstrap resampling procedure with 500 samples was used to internally validate the model, estimate optimism-corrected discrimination, and assess calibration.

### Model validation

Using the equation derived in the development cohort we calculated the predicted probability of developing type 2 diabetes for patients in the validation cohort. Model performance upon external validation was assessed for discrimination using Harrell’s measure of concordance for censored response variable and calibration.^19^

### Estimating risk-specific treatment effects

To estimate the risk-based treatment effect for metformin pharmacotherapy or the DPP lifestyle modification, we performed a risk-based heterogeneity of treatment effect analysis on the DPP.^20^ The applicability of the OLDW model to the DPP data was anticipated to be limited by: differences between predictor variable definitions and measurement within a trial context vs. EHR data, differences in the pattern of missingness between these contexts (i.e., there was essentially no data missingness in the DPP), differences in patient enrollment in the two settings and differences in outcome definition and ascertainment.^21^ Thus, we refit the OLDW model to the DPP, using the same variables and interaction terms. Consistent with methodological recommendations^22,23^, all 3 DPP arms were used, since research has shown that overfitting to a control arm can induce spurious heterogeneity of treatment effects.^24-26^ The treatment effect was then estimated by incorporating this linear predictor into a Cox proportional hazards model with the following terms: treatment (metformin or DPP lifestyle modification), the linear predictor of risk from the refitted model, and (potentially) an interaction between these to account for important changes in relative risk reduction across different levels of baseline risk. Based on a previous analysis,^8^ we anticipated a risk-by-treatment interaction with metformin pharmacotherapy and a consistent relative effect with the DPP lifestyle modification, but we examined interactions for both treatment arms. We also performed a sensitivity analysis, examining the risk-by-treatment interactions, stratifying the DPP by the OLDW model without any refitting, and examining the distribution of predicted effects using this model.

### Incorporation of Decision Support in Electronic Health Record

In order to facilitate use in clinical decision making, based on patient and provider focus groups and interviews, we implemented the model in two different ways: 1) a hard coded calculation in an Allscripts EHR; 2) a cloud-hosted SMART on FHIR^27^ app that can be incorporated into any EHR, leveraging interoperability standards recently promulgated by the US office of the National Coordinator of Health Information (ONC).

### IRB Approval

This study was reviewed and approved by the Tufts Health Sciences Institutional Review Board prior to accessing the deidentified data from the DPP and OLDW datasets.

## Results

Figure 1 shows the development of the derivation and validation OLDW datasets. Approximately 1.1 million people with prediabetes from the Northeast, South and West were included in the derivation cohort, and a similar number from the Midwest were included in the validation cohort. Characteristics of these cohorts are shown in Table 1.

**Figure 1.**
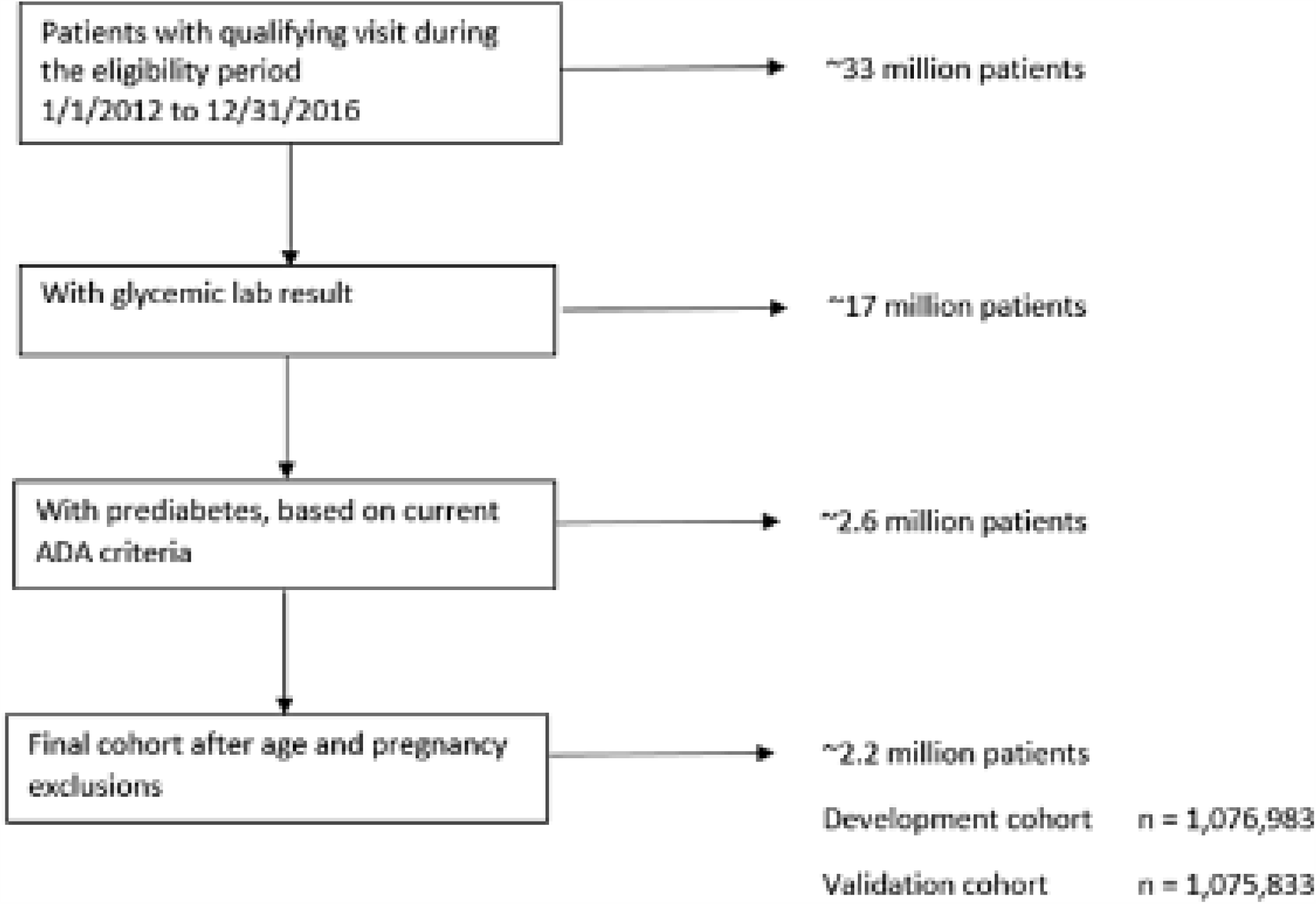
CONSORT Diagram for OLDW derivation and validation cohort.

### Model development and validation: Risk stratification

The coefficients for each of the variable and interaction terms included in the model are shown in Table 2. The optimism-corrected *c*-statistic on the derivation sample was 0.73. When the model was tested on the validation cohort, the *c*-statistic was slightly higher, 0.76. Calibration on the validation cohort was very good (Figure 2). Among the 268,959 patients in the lowest-risk quartile, the predicted diabetes rate was 3.1% (95% confidence interval, 3.0 to 3.2%), while the observed rate was 1.8% (1.7 to 1.9%); among the 268,958 patients in highest-risk quartile, the predicted diabetes rate was 19.2% (18.6 to 19.9%), while the observed rate was 19.6% (19.4 to 19.8%).

**Table 2.** Final model for incident diabetes.

|  | HR | Lower | Upper |  | HR | Lower | Upper |
| --- | --- | --- | --- | --- | --- | --- | --- |
| Age, per 10 years | 1.08 | 1.08 | 1.08 | *adjustments for missing data* |  |  |  |
| Female | 1.21 | 1.19 | 1.23 | Race(missing) vs White | 0.16 | 0.07 | 0.38 |
| Black vs white | 2.73 | 1.32 | 5.64 | Smoking(missing) vs never | 1.08 | 1.06 | 1.11 |
| Asian vs white | 0.01 | 0.00 | 0.02 | A1c(missing) | 0.75 | 0.74 | 0.77 |
| Current smoker vs never | 1.22 | 1.19 | 1.24 | FPG(missing) | 1.03 | 0.99 | 1.07 |
| Former smoker vs never | 1.11 | 1.09 | 1.13 | Triglycerides(missing) | 1.08 | 1.03 | 1.12 |
| HTN | 1.23 | 1.21 | 1.25 | BMI(missing) | 1.22 | 1.18 | 1.26 |
| A1c, per 0.1% | 1.24 | 1.18 | 1.29 | SBP(missing) | 1.22 | 1.17 | 1.26 |
| FPG, per 10 mg/dL | 1.29 | 1.29 | 1.29 | HDL(missing) | 1.23 | 1.17 | 1.28 |
| Triglycerides, per 10 mg/dL | 1.01 | 1.01 | 1.02 | AA*BMI(missing) | 0.97 | 0.91 | 1.03 |
| BMI, per 5 units | 1.24 | 1.24 | 1.24 | AA*A1c(missing) | 1.56 | 1.48 | 1.64 |
| SBP, per 20 mmHG | 1.05 | 1.05 | 1.05 | Asian*BMI(missing) | 0.77 | 0.70 | 0.84 |
| HDL, per 10 mg/dL | 0.85 | 0.85 | 0.85 | Asian*A1c(missing) | 2.03 | 1.85 | 2.23 |
| Black *BMI | 0.98 | 0.98 | 0.99 | Race(missing)*BMI | 0.99 | 0.99 | 1.00 |
| Black *A1c | 0.95 | 0.84 | 1.07 | Race(missing)*BMI(missing) | 0.82 | 0.77 | 0.87 |
| Asian*BMI | 1.00 | 0.99 | 1.01 | Race(missing)*A1c | 1.40 | 1.22 | 1.61 |
| Asian*A1c | 2.32 | 1.91 | 2.83 | Race(missing)*A1c(missing) | 1.46 | 1.37 | 1.55 |

Baseline hazard at 5 years (S0):
1 year = 0.02470
2 years = 0.04757
3 years = 0.07044

**Figure 2.**
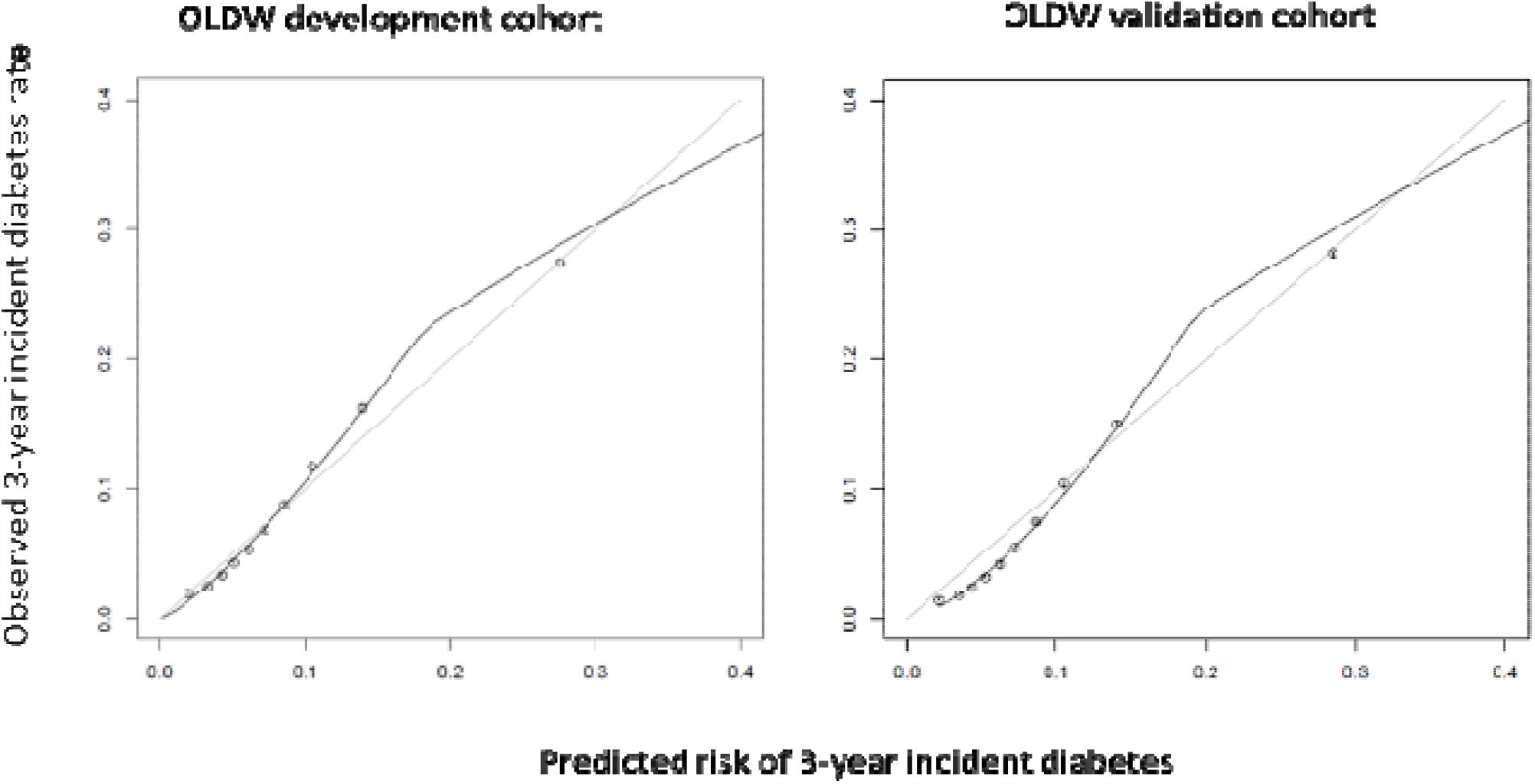
Calibration Curves. The figure on the left depicts the observed versus the predicted 3-year rate of developing diabetes in the 1.1 million patients in the derivation cohort (the Northeast, South and West regions) divided into equal sized tenths. The figure on the risk depicts the observed versus the predicted 3-year rate of developing diabetes in the 1.1 million patients in the validation cohort (the Midwest).

### Calculation of relative treatment effects in the DPP Study

Prior work demonstrated a consistent relative treatment effect across risk groups with the DPP lifestyle modification and an increasing relative effect with progressively higher risk for metformin pharmacotherapy.^8^ Using the OLDW model refit to the DPP data (Appendix Table 2; *c*-statistic 0.719), we confirmed the absence of a treatment-by-risk interaction for lifestyle modification (p for interaction = 0.68); thus, we applied a constant relative risk reduction in the prediction model (HR = 0.43; 95% CI: 0.35 – 0.53) to estimate the diabetes outcome with lifestyle modification. We also confirmed the presence of a treatment-by-risk interaction with metformin pharmacotherapy (p for interaction= 0.003; using the continuous risk on the logit scale): low-risk patients had outcomes with metformin that were similar to usual care (in lowest risk quarter, observed HR = 1.1; 95% CI: 0.61 to 2.0), and high-risk patients have outcomes with metformin that were similar to the DPP lifestyle modification (in highest risk quarter, observed HR = 0.45; 95% CI: 0.35 to 0.59). Figure 3 shows observed and predicted benefits across quartiles for the DPP, for both lifestyle and metformin. A look-up table showing the relative risk reduction with metformin for each level of risk is shown in Appendix Table 3, truncated at a low value of 0% (no harm or benefit) and a high value of 60%.

**Figure 3.**
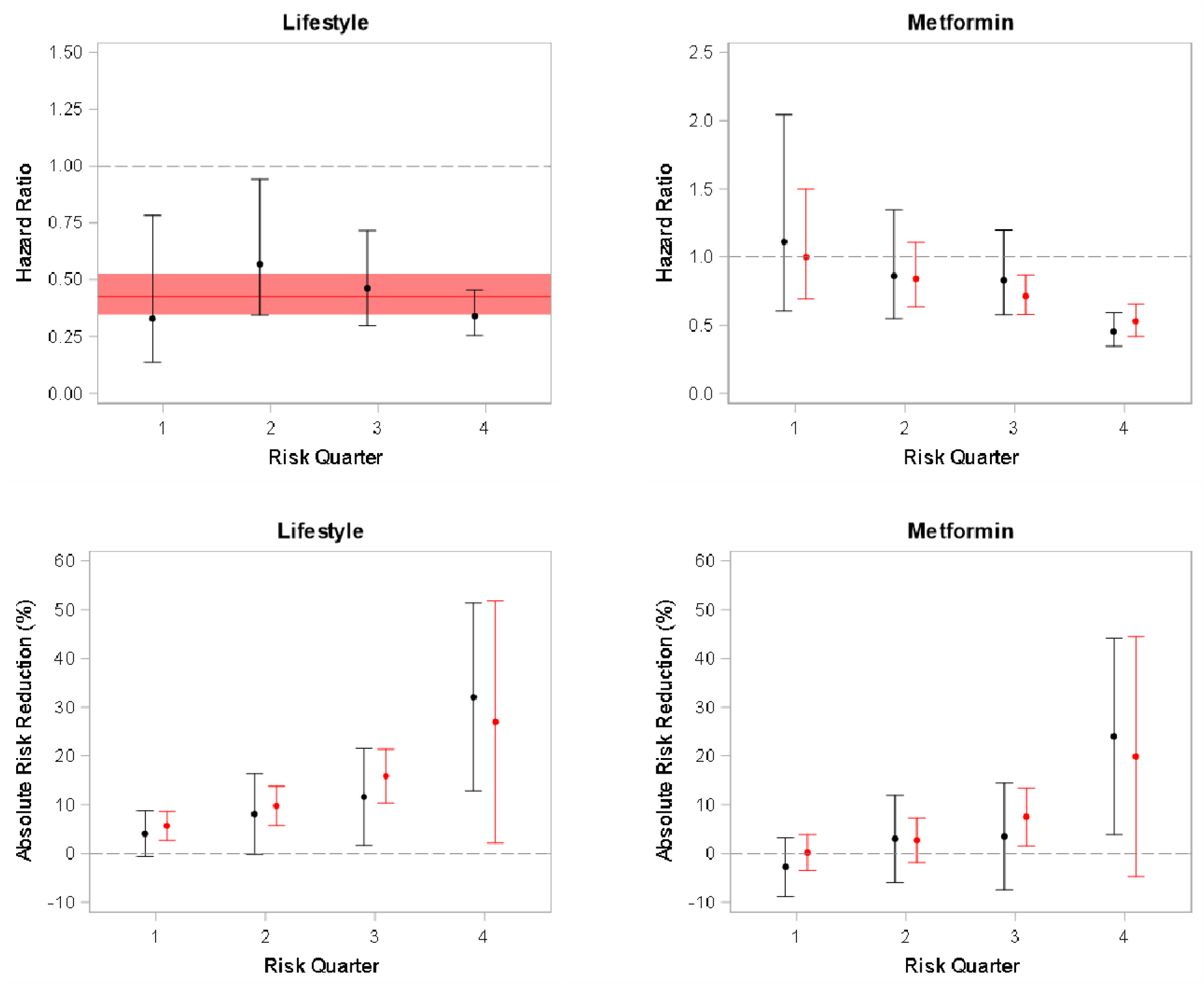
Observed and predicted treatment effects in the DPP Study Across Risk Groups. Black dot and bar is observed treatment effect. Red dot and bar is predicted treatment effect. Figure 3 depicts the observed treatment effects (black dots) in patients in the DPP Study when patients are stratified into quarters based on their predicted risk for the DPP lifestyle modification intervention (left) and for metformin (right). Predicted effects across risk groups are shown in red. The top set of graphs displays relative effects and shows a consistency of effects across risk groups for lifestyle modification but heterogeneous treatment effects for metformin (p=0.003). The bottom graphs show effects on the absolute risk difference scale, which shows increasing benefits for higher risk patients for both interventions.

### Distribution of risks and benefits in OLDW

The overall average 3-year predicted risk of developing diabetes for patients in the validation OLDW cohort was 9.0%, 3.9%, and 6.0%, with usual care, the DPP lifestyle diabetes and metformin respectively. Predictions for the median patient in each quartile using the final model are shown in Table 2. For lifestyle modification, 53% of the total preventable cases of diabetes could be prevented by treating the 25% of patients at highest risk; 76% by treating the 50% at highest risk and 91% by treating the 75% at highest risk. For metformin therapy, 73% of the total preventable cases could be prevented by treating the 25% of patients at highest risk; 93% by treating the 50% at highest risk, and 100% by treating the 75% at highest risk.

### Sensitivity analysis

Direct application of the OLDW model (not refit) on the DPP showed a moderately diminished discrimination (*c*-statistic = 0.68). There was no risk-by treatment interaction with lifestyle (p = 0.69). The risk-by-treatment interaction with metformin was qualitatively similar to that with the refit model (p = 0.08), and the distribution of predicted benefits with this model was also similar. For lifestyle modification, 53% of the total cases of preventable diabetes could be prevented by treating the 25% of patients at highest risk; 76% by treating the 50% at highest risk. For metformin therapy, 65% of the total cases of preventable diabetes could be prevented by treating the 25% of patients at highest risk; 86% by treating the 50% at highest risk.

### Implementation of the final model

Figure 4 shows the user interface of the SMART app in an EHR. Predictions are generated automatically based on the data available and retrieved from the patient’s record, using appropriate indicators in the model for missingness where necessary.

**Figure 4.**
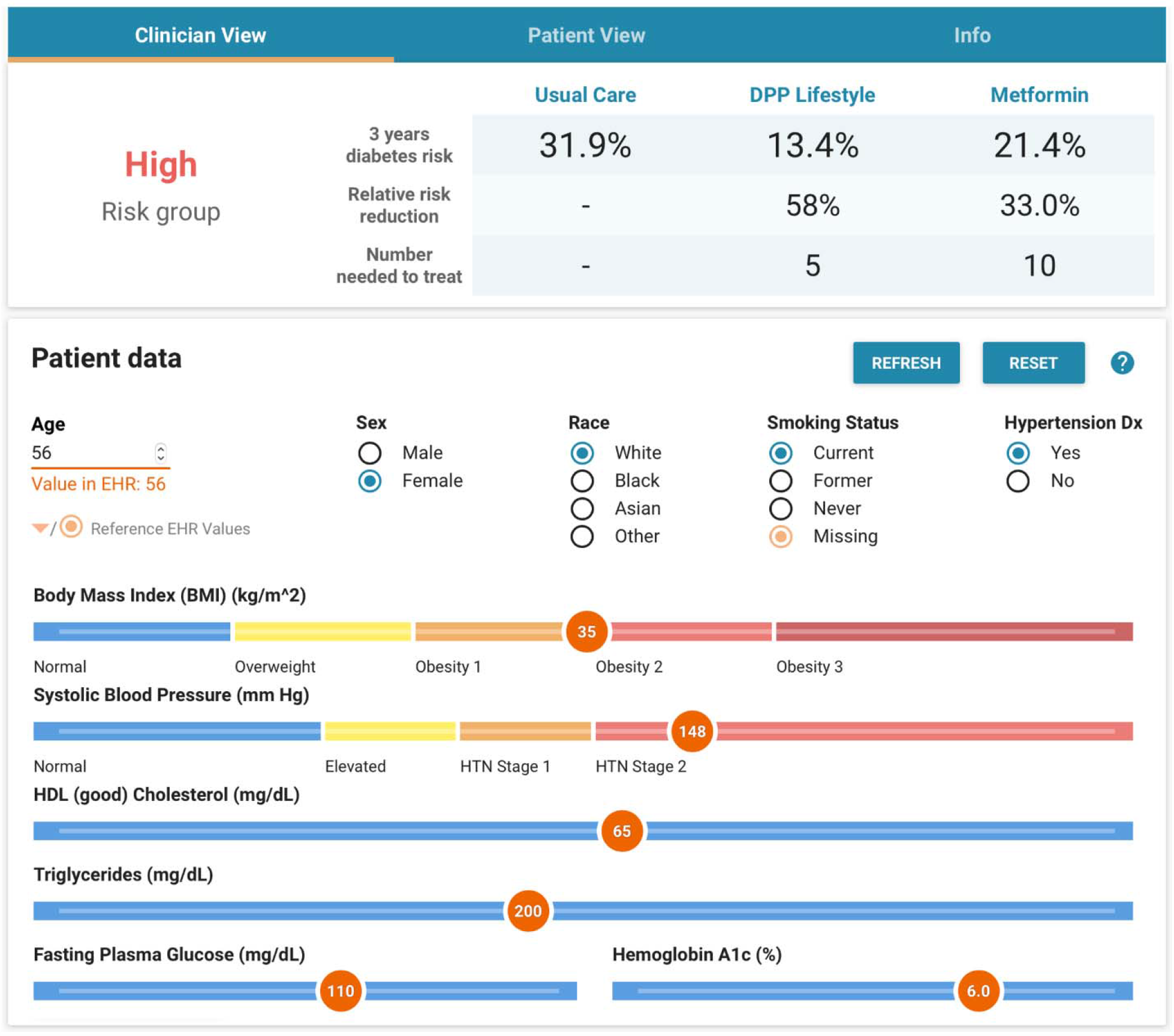
User interface for decision support tool.

## Discussion

We present an EHR-compatible model to predict diabetes onset, using 11 variables routinely collected in clinical practice. A major strength of the risk model is that it was derived on the OLDW, which reflects people with prediabetes defined by the most commonly used ADA criteria, from heterogeneous EHRs and more than 30 US healthcare systems. The risk model derived in 3 US Census regions performed very well in a geographically distinct cohort. Compatible risk-specific estimates of treatment effect were then obtained directly from the DPP. By prioritizing care based on the risk of diabetes, this “hybrid” model might help optimize the efficiency of diabetes prevention: Treating just the highest-risk half of people with prediabetes would capture 77% of the benefit of population-wide lifestyle modification or 93% of the benefit of population-wide metformin pharmacotherapy. This is important because lifestyle programs are resource-intensive and require a high level of commitment from the patient. Pharmacotherapy is not without adverse effects, and over-treatment should be avoided, especially in low-risk patients who do not appear to benefit.

The issue of how to address prediabetes has grown in importance as broader diabetes screening has been recommended and promoted.^13,28^ For every patient with diabetes identified, screening identifies 6 patients with prediabetes; health systems are thus confronted with a growing number of patients who have prediabetes, without the capacity to treat everybody, reserving limited resources to improving cardiometabolic control for patients with diabetes. While the ADA has lowered the A1c and FG thresholds to define prediabetes,^9,29^ some have argued that the value of medicalizing prediabetes and defining an ever-growing proportion of the population as diseased is of dubious value.^5^ Most patients who are classified as prediabetic do not develop diabetes even in a decade, and risks of developing end organ damage are low for those developing diabetes later in life.^30^ Risk stratification offers an approach that promises more focused resources specifically on those who are likely to benefit. While our prior research results provided proof-of-concept that risk stratification could support providers and health systems prioritize these patients,^8^ the present EHR-compatible model is designed to be used at point of care, and it has been incorporated into the EHRs at several locations in the US.

A longstanding concern regarding limitations of randomized clinical trial results is that they might not be applicable when there is non-random selection into the trial and when treatment effects are heterogeneous^31^. Here, for example, we found that the “real world” at-risk population was at substantially lower overall risk than patients included in the DPP, and that treatment effects were risk-dependent. The lower overall risk in the OLDW cohort is the result of multiple factors, including: 1) different inclusion criteria for the DPP (including a high BMI and elevated 2-hour glucose after a 75-gram glucose load); 2) differences in the distribution of risk variables; 3) different outcome ascertainment, which is substantially more rigorous in the trial setting. Cross-design synthesis has been proposed as a means of addressing the potential problems of external validity of trial evidence by combining the strengths of both designs—observational designs to capture the full range of patients and randomized trials for unbiased treatment effects^32,33^.

While several different methods for cross-design synthesis have been proposed^34,35^, all approaches depend on the ability to adjust results based on patient characteristics across designs. A seldom-discussed barrier is that variable definitions and ascertainment can differ considerably between clinical trial data and routinely collected observational data. Our approach was designed to address these barriers in a pragmatic way, by estimating risk-specific treatment effects in the clinical trial using the same set of variables as used in the observational risk model. This approach was driven in part by our novel aim, to predict effects in patients in clinical care, based on individual patient characteristics, rather than estimating average treatment effects in broad target populations.

A related issue that has received limited attention is how to deploy clinical prediction models in an EHR. There is a proliferation of clinical prediction models; use of routinely collected EHR data to automatically generate individual patient predictions is an appealing approach to disseminate these into the clinic. However, most published clinical prediction models are developed on research cohorts or clinical trials. Predictor variables collected in a trial are not consistently and rigorously captured in the EHR. Recent work has highlighted that heterogeneity in predictor measurement across different settings can substantially degrade model performance.^21,36^ Finally, use of trial or registry data cannot yield a model robust to missing values in the EHR database used for clinical prediction, since the pattern of missingness present across research and EHR environments is expected to differ. The usual approaches addressing potential bias arising from missingness (e.g., multiple imputation) are not designed to cope with missingness in variables used to generate predictions. These issues guided our decision to derive separate models in the EHR and trial setting, using a common set of variables that were well ascertained in both settings.

### Limitations

The methods we used for “cross-walking” between the two very different types of data (trial and EHR real world data) potentially introduce estimation error. Ideally, individualized treatment effects would be estimated on databases that combine the advantages of these different data sources: unbiased effect estimates through randomization; meticulous outcome ascertainment; consistency of predictors across derivation and implementation populations, and large, heterogeneous populations. Improving the quality of data collection in routine care and integrating randomized trials into routine care^37-39^ may narrow the gap between trial and “real-world” data. Despite these limitations, we obtained qualitatively consistent risk-stratified results in the DPP regardless of which risk model was used: consistency of relative treatment effects of lifestyle modification across all levels of risk and heterogeneous relative treatment effects with metformin, with much stronger relative effects in higher-risk patients.

## Conclusion

While the number of people in the US who have prediabetes and qualify for diabetes prevention programs could potentially overwhelm health care systems, these patients have substantial variation in their risk of developing diabetes and in their likelihood of benefiting from prevention therapies. Incorporation of a tool into the EHR to support automated risk stratification of patients in routine clinical care—by predicting individualized benefits—can support shared decision-making and prioritize those patients who are most likely to benefit, where capacity might be limited.

## Supporting information

STROBE Checklist

DKent COI Form

## Data Availability

The data that support the findings of this study are available from OptumLabs but restrictions apply to the availability of these data, which were used under license for the current study, and so are not publicly available. Data are however available from the authors upon reasonable request and with permission of OptumLabs.

## Funding/Acknowledgements

This work was supported by a Patient-Centered Outcomes Research Institute (PCORI) contract (DI-1604-35234). David Kent is the guarantor for this manuscript.

