## Appendix for "An Electronic Health Record Compatible Model to Predict Personalized Treatment Effects from the Diabetes Prevention Program: A Cross-Evidence Synthesis Approach Using Clinical Trial and Real World Data"

**Appendix Table 1. Risk factor definitions and diagnosis codes**

Eligibility Criteria:

- Office visit (see definitions below) between 1/1/2012 and 12/31/2016 (study dates)
- Age 25-75 at the time of encounter
- On or prior to this office visit (within 12 months) patient has one of the following pre-diabetes criteria:
  - Pre-diabetes A1C: 5.7-6.4%
  - Pre-diabetes fasting glucose (or glucose measured on same day as lipid panel): 100-125 mg/dL
- Prior to this office visit patient has at least one documented encounter (ie criteria to define enrollment in system)
- **Exclude** any patients with type 1 or type 2 diabetes by any of the following criteria on or prior to this office visit:
  - Diabetes diagnosis: ICD-09 250.* / ICD-10 E10.* or E11.*
  - Diabetes related pharmacotherapy or procedure (ie HEDIS criteria)
  - A1C: greater than 6.4
  - Fasting glucose (or glucose measured on same day as lipid panel): greater than 125 mg/dL
  - Random glucose >=200 mg/dL on 2 occasions (within 3 months of each other)
  - 2 hour glucose > 199 mg/dL
- Also exclude patients with any of the following documented conditions:
  - pregnant within 24 months of index visit

Outcome definition:

- **Time to first encounter** (from index office visit) meeting type 2 diabetes by any of the following criteria:
  - Diabetes diagnosis: ICD-09 250.* / ICD-10 E10.* or E11.*
  - Diabetes related pharmacotherapy or procedure (by HEDIS criteria)
  - A1C: greater than 6.4 (requires confirmation by HEDIS criteria or additional lab)
  - Fasting glucose (or glucose measured on same day as lipid panel): greater than 125 mg/dL (requires confirmation by HEDIS criteria or additional lab)
  - 2 hour > 199 (requires confirmation by HEDIS criteria or additional lab)
- Censor follow up for patients who do not meet outcome definition on the earliest occurrence of any of the following:
  - time of death
  - last encounter or end of enrollment in system
  - initiation of diabetes medication

**Office visit definitions used for OLDW cohort:**

Has an office visit during study period defined as any of the following:

- Patient encounter (variable name: “INTERACTION_TYPE”) categorized as “Office or clinic patient”
- Patient encounter with one of the following procedure codes:

| CPT/HCPCS Codes | Description |
| --- | --- |
| 99201–99205, 99211–99215 | Evaluation & Management Office Visit |
| 99241–99245 | Evaluation & Management Office Consultation |
| 99381–99387, 99391–99397 | Evaluation & Management Preventive Visit |
| 99401–99404 | Preventative Medicine: Individual Counseling Visit |
| 99411–99412 | Preventative Medicine: Group Counseling Visit |
| 99420, 99429 | Other Preventive Medicine Services |
| G0402 | Initial Preventive Physical Examination (“Welcome to Medicare” Visit) |
| G0438, G0439 | Medicare Annual Wellness Visit |
| G0463 | Hospital outpatient clinic visit for assessment and management of a patient |
| T1015 | Clinic visit/encounter, all inclusive |

**Appendix Table 2.** **Refit model using complete case DPP data for estimating metformin treatment benefit**

|  | HR | Lower 95% CI | Upper 95% CI | z | p |
| --- | --- | --- | --- | --- | --- |
| Age | 0.991 | 0.980 | 1.002 | -1.668 | 0.095 |
| Female sex | 1.061 | 0.874 | 1.287 | 0.594 | 0.553 |
| Black race | 9.812 | 0.847 | 113.672 | 1.827 | 0.068 |
| Other race | 3.815 | 0.011 | 1362.586 | 0.446 | 0.655 |
| Current smoker | 1.223 | 0.944 | 1.584 | 1.523 | 0.128 |
| Former smoker | 0.954 | 0.798 | 1.141 | -0.516 | 0.606 |
| Hypertension | 1.092 | 0.898 | 1.328 | 0.882 | 0.378 |
| A1c | 2.383 | 1.925 | 2.950 | 7.978 | <0.001 |
| FPG | 1.068 | 1.058 | 1.079 | 12.877 | <0.001 |
| Triglycerides | 1.002 | 1.001 | 1.003 | 3.783 | <0.001 |
| BMI | 1.008 | 0.991 | 1.026 | 0.925 | 0.355 |
| SBP | 1.003 | 0.997 | 1.010 | 0.954 | 0.340 |
| HDL | 0.995 | 0.986 | 1.004 | -1.163 | 0.245 |
| Black*BMI | 0.992 | 0.959 | 1.026 | -0.456 | 0.648 |
| Black*A1c | 0.697 | 0.491 | 0.989 | -2.024 | 0.043 |
| Other race*BMI | 1.017 | 0.952 | 1.086 | 0.492 | 0.623 |
| Other race*A1c | 0.723 | 0.285 | 1.830 | -0.685 | 0.493 |
| *C*-statistic | | 0.719 |  |  |  |
| Optimism corrected c-statistic | | 0.710 |  |  |  |
| Calibration slope (bootstrap) | | 0.951 |  |  |  |
| Baseline hazard at 3 years | | 0.2016 |  |  |  |

**Appendix Table 3. Metformin relative risk reduction look-up table**

| **3-year risk (usual care)** | **HR** | **RRR** | **3-year risk (metformin)** | **ARR** | **NNT** |  |
| --- | --- | --- | --- | --- | --- | --- |
| 1% | 1.00 | 0.00 | 1.0% | 0.0% | - | HR = hazard ratio |
| 2% | 1.00 | 0.00 | 2.0% | 0.0% | - | RRR= relative risk reduction |
| 3% | 1.00 | 0.00 | 3.0% | 0.0% | - | ARR = absolute risk reduction |
| 4% | 1.00 | 0.00 | 4.0% | 0.0% | - | NNT= number needed to treat |
| 5% | 1.00 | 0.00 | 5.0% | 0.0% | - |  |
| 6% | 1.00 | 0.00 | 6.0% | 0.0% | - |  |
| 7% | 1.00 | 0.00 | 7.0% | 0.0% | - |  |
| 8% | 1.00 | 0.00 | 8.0% | 0.0% | - |  |
| 9% | 0.99 | 0.01 | 8.9% | 0.1% | 1558 |  |
| 10% | 0.96 | 0.04 | 9.6% | 0.4% | 241 |  |
| 11% | 0.92 | 0.08 | 10.1% | 0.9% | 110 |  |
| 12% | 0.89 | 0.11 | 10.7% | 1.3% | 79 |  |
| 13% | 0.86 | 0.14 | 11.2% | 1.8% | 57 |  |
| 14% | 0.84 | 0.16 | 11.8% | 2.2% | 45 |  |
| 15% | 0.82 | 0.18 | 12.3% | 2.7% | 37 |  |
| 16% | 0.80 | 0.20 | 12.8% | 3.2% | 31 |  |
| 17% | 0.78 | 0.22 | 13.2% | 3.8% | 27 |  |
| 18% | 0.76 | 0.24 | 13.6% | 4.4% | 23 |  |
| 19% | 0.74 | 0.26 | 14.1% | 4.9% | 21 |  |
| 20% | 0.73 | 0.27 | 14.6% | 5.4% | 19 |  |
| 21% | 0.71 | 0.29 | 15.0% | 6.0% | 17 |  |
| 22% | 0.70 | 0.30 | 15.4% | 6.6% | 16 |  |
| 23% | 0.69 | 0.31 | 15.9% | 7.1% | 14 |  |
| 24% | 0.68 | 0.32 | 16.3% | 7.7% | 13 |  |
| 25% | 0.67 | 0.33 | 16.7% | 8.3% | 12 |  |
| 26% | 0.65 | 0.35 | 17.0% | 9.0% | 12 |  |
| 27% | 0.65 | 0.35 | 17.5% | 9.5% | 11 |  |
| 28% | 0.64 | 0.36 | 17.9% | 10.1% | 10 |  |
| 29% | 0.63 | 0.37 | 18.2% | 10.8% | 10 |  |
| 30% | 0.62 | 0.38 | 18.5% | 11.5% | 9 |  |
| 31% | 0.61 | 0.39 | 19.0% | 12.0% | 9 |  |
| 32% | 0.60 | 0.40 | 19.3% | 12.7% | 8 |  |
| 33% | 0.60 | 0.40 | 19.7% | 13.3% | 8 |  |
| 34% | 0.59 | 0.41 | 19.9% | 14.1% | 8 |  |
| 35% | 0.58 | 0.42 | 20.4% | 14.6% | 7 |  |
| 36% | 0.57 | 0.43 | 20.6% | 15.4% | 7 |  |
| 37% | 0.57 | 0.43 | 21.0% | 16.0% | 7 |  |
| 38% | 0.56 | 0.44 | 21.4% | 16.6% | 7 |  |
| 39% | 0.55 | 0.45 | 21.6% | 17.4% | 6 |  |
| 40% | 0.55 | 0.45 | 21.9% | 18.1% | 6 |  |
| 41% | 0.54 | 0.46 | 22.3% | 18.7% | 6 |  |
| 42% | 0.53 | 0.47 | 22.5% | 19.5% | 6 |  |
| 43% | 0.53 | 0.47 | 22.8% | 20.2% | 5 |  |
| 44% | 0.53 | 0.47 | 23.1% | 20.9% | 5 |  |
| 45% | 0.52 | 0.48 | 23.5% | 21.5% | 5 |  |
| 46% | 0.51 | 0.49 | 23.6% | 22.4% | 5 |  |
| 47% | 0.51 | 0.49 | 23.9% | 23.1% | 5 |  |
| 48% | 0.50 | 0.50 | 24.2% | 23.8% | 5 |  |
| 49% | 0.50 | 0.50 | 24.5% | 24.5% | 5 |  |
| 50% | 0.50 | 0.50 | 24.8% | 25.2% | 4 |  |
| 51% | 0.49 | 0.51 | 25.1% | 25.9% | 4 |  |
| 52% | 0.48 | 0.52 | 25.2% | 26.8% | 4 |  |
| 53% | 0.48 | 0.52 | 25.4% | 27.6% | 4 |  |
| 54% | 0.48 | 0.52 | 25.7% | 28.3% | 4 |  |
| 55% | 0.47 | 0.53 | 26.0% | 29.0% | 4 |  |
| 56% | 0.47 | 0.53 | 26.2% | 29.8% | 4 |  |
| 57% | 0.46 | 0.54 | 26.5% | 30.5% | 4 |  |
| 58% | 0.46 | 0.54 | 26.7% | 31.3% | 4 |  |
| 59% | 0.46 | 0.54 | 26.9% | 32.1% | 4 | **Optum cohort 90th percentile of risk (59%)** |
| 60% | 0.45 | 0.55 | 27.0% | 33.0% | 4 |  |
| 61% | 0.45 | 0.55 | 27.2% | 33.8% | 3 |  |
| 62% | 0.44 | 0.56 | 27.4% | 34.6% | 3 |  |
| 63% | 0.44 | 0.56 | 27.6% | 35.4% | 3 |  |
| 64% | 0.43 | 0.57 | 27.8% | 36.2% | 3 |  |
| 65% | 0.43 | 0.57 | 28.0% | 37.0% | 3 |  |
| 66% | 0.43 | 0.57 | 28.2% | 37.8% | 3 |  |
| 67% | 0.42 | 0.58 | 28.4% | 38.6% | 3 |  |
| 68% | 0.42 | 0.58 | 28.6% | 39.4% | 3 |  |
| 69% | 0.42 | 0.58 | 28.8% | 40.2% | 3 |  |
| 70% | 0.41 | 0.59 | 29.0% | 41.0% | 3 |  |
| 71% | 0.41 | 0.59 | 28.9% | 42.1% | 3 | **Optum cohort 95th percentile of risk (71%)** |
| 72% | 0.40 | 0.60 | 29.1% | 42.9% | 3 |  |
| 73% | 0.40 | 0.60 | 29.2% | 43.8% | 3 | **RRR truncated at 60%** |
| 74% | 0.40 | 0.60 | 29.6% | 44.4% | 3 |  |
| 75% | 0.40 | 0.60 | 30.0% | 45.0% | 3 |  |
| 76% | 0.40 | 0.60 | 30.4% | 45.6% | 3 |  |
| 77% | 0.40 | 0.60 | 30.8% | 46.2% | 3 |  |
| 78% | 0.40 | 0.60 | 31.2% | 46.8% | 3 |  |
| 79% | 0.40 | 0.60 | 31.6% | 47.4% | 3 |  |
| 80% | 0.40 | 0.60 | 32.0% | 48.0% | 3 |  |
| 81% | 0.40 | 0.60 | 32.4% | 48.6% | 3 |  |
| 82% | 0.40 | 0.60 | 32.8% | 49.2% | 3 |  |
| 83% | 0.40 | 0.60 | 33.2% | 49.8% | 3 |  |
| 84% | 0.40 | 0.60 | 33.6% | 50.4% | 2 |  |
| 85% | 0.40 | 0.60 | 34.0% | 51.0% | 2 |  |
| 86% | 0.40 | 0.60 | 34.4% | 51.6% | 2 |  |
| 87% | 0.40 | 0.60 | 34.8% | 52.2% | 2 |  |
| 88% | 0.40 | 0.60 | 35.2% | 52.8% | 2 |  |
| 89% | 0.40 | 0.60 | 35.6% | 53.4% | 2 |  |
| 90% | 0.40 | 0.60 | 36.0% | 54.0% | 2 |  |
| 91% | 0.40 | 0.60 | 36.4% | 54.6% | 2 |  |
| 92% | 0.40 | 0.60 | 36.8% | 55.2% | 2 |  |
| 93% | 0.40 | 0.60 | 37.2% | 55.8% | 2 |  |
| 94% | 0.40 | 0.60 | 37.6% | 56.4% | 2 |  |
| 95% | 0.40 | 0.60 | 38.0% | 57.0% | 2 |  |
| 96% | 0.40 | 0.60 | 38.4% | 57.6% | 2 |  |
| 97% | 0.40 | 0.60 | 38.8% | 58.2% | 2 |  |
| 98% | 0.40 | 0.60 | 39.2% | 58.8% | 2 |  |
| 99% | 0.40 | 0.60 | 39.6% | 59.4% | 2 |  |
| 100% | 0.40 | 0.60 | 40.0% | 60.0% | 2 |  |
